# Behavioral Effects in Disorders of Consciousness Following Transcranial Direct Current Stimulation: A Systematic Review and Individual Patient Data Meta-analysis of Randomized Clinical Trials

**DOI:** 10.1101/2022.05.08.22274809

**Authors:** Zeyu Xu, Ruizhe Zheng, Tiantong Xia, Zengxin Qi, Di Zang, Zhe Wang, Xuehai Wu

**Author notes:** **Correspondence author:** Xuehai Wu,.

## Abstract

**Background:** In patients with Disorders of Consciousness (DoC), transcranial direct current stimulation (tDCS) was a promising intervention for it. However, uncertainties remain about the treatment effect and the optimal treatment strategy of the tDCS in the DoC.

**Objective:** In this meta-analysis of individual patient data (IPD), we assess whether utilizing tDCS as a treatment in DoC could improve patients’ behavioral performance and whether patient characteristics or tDCS protocol could modify the treatment effect.

**Methods:** We searched PubMed, Embase, and the Cochrane Central Register of Controlled Trials through April 7, 2022, using the terms “persistent vegetative state,” “minimally conscious state,” “disorder of consciousness,” or “unresponsive wakefulness syndrome,” and “transcranial direct current stimulation” to identify Randomized Controlled Trials (RCTs) in English-language publication. Studies were eligible for inclusion if they reported pre- and post-tDCS Coma Recovery Scale-Revised (CRS-R) scores. From the included studies, any patients who had incomplete data were excluded. We performed a meta-analysis to assess the treatment effect of the tDCS compared with sham control. Additionally, a subgroup analysis was performed to determine whether patients’ baseline characteristics could modify the treatment effect and the optimal tDCS protocol.

**Results:** We identified 145 papers, eight trials (including 181 patients) were finally included in the analysis, and one individual data were excluded because of incompletion. Our meta-analysis demonstrated a mean difference change in the CRS-R score of 0.89 (95% CI, 0.17-1.61) between tDCS and sham-control, favoring tDCS. The subgroup analysis showed that patients who were male or in minimally conscious state (MCS) were associated with a greater improvement in CRS-R score and that adopting 5 or more sessions of tDCS protocol might have a better treatment effect than just one session.

**Conclusion:** tDCS can improve the behavioral performance of DoC patients. However, heterogeneity existed within the patients’ baseline condition and the stimulation protocol. There should be more exploration of the optimal tDCS protocol and the most beneficial patient group based on the mechanism of tDCS in the future.

## Introduction

Disorders of Consciousness (DoC) is defined as alterations in arousal or awareness, often caused by cardiac arrest, traumatic brain injury (TBI), intracerebral hemorrhage, and so on[1]. With the development of technology in neurological intensive care and neurosurgery, more and more patients survived these acute diseases. Still, the number of patients who can not regain consciousness after these diseases have evolved either. In the past several decades, a lot of work has been done regarding the diagnosis and prognosis of DoC, making our understanding of DoC advanced. However, therapeutical intervention option in DoC was scarce. The 2018 American practice guidelines for DoC patients [2] only recommend amantadine for treatment. Even though many brain stimulation therapy emerged in recent years but are still in research, including transcranial direct current stimulation (tDCS), transcranial magnetic stimulation (TMS), deep brain stimulation, and so on.

Transcranial direct current stimulation (tDCS) is a non-invasive brain stimulation technique that modulates the activity of targeted brain regions by using a low-intensity direct current (usually 1–2 mA) between two electrodes (an anode and a cathode) placed on the scalp. [3] Among recent studies, tDCS had shown promising results in DoC. A meta-analysis of non-invasive stimulation treatment in DoC patients demonstrates that tDCS over the dorsolateral prefrontal cortex (DLPFC) is able to improve the Coma Recovery Score –Revised (CRS-R) score. [4] Nevertheless, five recent randomized clinical trials (RCT) reported no treatment effect between treatment and control.[5-9] Possible reasons for these heterogeneous results are related to the different tDCS protocols, such as session numbers and duration, current intensity, stimulation target, and so on. For example, Zhang et al. [10] showed that tDCS over the DLPFC improved behavioral responsiveness in patients, whereas Martens et al. found no improvement over the primary motor cortex (M1). Patients’ baseline characteristics also play an important role, and several studies had revealed a better treatment effect in a patient with a baseline diagnosis as minimally conscious states (MCS) than in persistent vegetative state (PVS).[11, 12] In sum, uncertainties still remain about the treatment effect of the tDCS in DoC and the optimal treatment strategy.

Therefore, we aim to perform an individual patient data meta-analysis from randomized clinical trials to assess the treatment effect of tDCS. We also sought to assess whether patient characteristics modify the treatment effect and find the optimal tDCS protocol.

## Methods

### Search strategy and selection criteria

This systematic review and individual patient data meta-analysis is conducted in accordance with the Preferred Reporting Items for Systematic Reviews and Meta-Analyses of individual participant data (PRISMA-IPD) guidelines[13]. We searched PubMed, Embase, and the Cochrane Central Register of Controlled Trials through April 7, 2022, using the terms “persistent vegetative state,” “minimally conscious state,” “disorder of consciousness,” or “unresponsive wakefulness syndrome,” and “transcranial direct current stimulation” to identify randomized controlled trials of studies utilizing tDCS as an intervention for DoC patients. The full search strategy is described in **Appendix 1** in the Supplement. Studies were included if: (1) Studies recruited patients diagnosed with MCS, PVS, or UWS, and (2) used tDCS as an intervention, and (3) with a sham stimulation as the control; and (4) pre- and post-tDCS Coma Recovery Scale-Revised (CRS-R) scores were used to measure the recovery in DoC patients as the outcomes; and (5) randomized controlled trials in either cross-over or parallel design; and (6) the authors provided individual patient data. Studies were published as the conference abstract, narrative or systematic reviews, or in books were excluded. Additionally, we excluded papers that were not randomized controlled trials or observational studies. Articles that were not accessible in English were also excluded. Two authors (ZYX and RZZ) independently identified studies meeting the inclusion criteria and excluded unrelated studies. Conflicts were resolved by consensus.

### Data collection and management

The following study-level data were extracted: first author, year of publication, study design, numbers of included patients, tDCS protocol that includes the number of sessions, current intensity, stimulation duration, stimulation site, and any adverse effects reported. Additionally, the following patient-level data were also extracted: age, gender, diagnosis, etiology of injury, time from injury to tDCS intervention, and CRS-R score at baseline and after the intervention. After individual patient data were collected, variables were transformed when possible to create a uniform database.

The risk of bias in each trial was assessed by two authors independently using the Cochrane Risk of Bias Tool.[14] Two authors (ZYX and RZZ) checked the IPD for all patients. Any suspected duplicated patients were discussed and decided whether they should be excluded or not.

### Outcome

The outcome was the behavioral effects of the tDCS treatment, measured via the absolute change in CRS-R score between pre-tDCS baseline score and post-tDCS score at the time point after finishing all sessions of tDCS treatment. CRS-R score is the most commonly used validated behavioral scale for DoC patients[15] and received strong recommendations from recent guidelines. [2, 16] It has six sub-scale, measured functions of auditory, visual, motor, verbal, communication, and arousal, respectively. The total CRS-R score ranges from 0 to 23, appearing capable of differentiating patients in an MCS from those in a VS.[17]

### Data analysis

Statistical analysis for outcome of interest was performed with IPD, according to the intention-to-treatment principle. The analysis involved both one-stage and two-stage methods for the outcome; In the one-stage method, a generalized linear mixed-effects model was conducted to analyze all trials simultaneously. In the two-stage method, we first analyzed each trial separately and then used a random-effects meta-analysis model to account for variability between trials and combine them. Adjusted analyses were performed to account for potential baseline incomparability. Adjustments were planned for the following prespecified covariates: age, sex, baseline CRS-R score, etiology of injury, time from injury to tDCS intervention. Results are reported as the mean difference in treatment effects between treatment and control with accompanying 95% CIs. Heterogeneity was evaluated by I^2^, and between-study variance (τ^2^). We performed the prespecified analysis in the following subgroup: age (non-elder with age <65 years vs. elder with age ≥65 years); gender (male vs. female); diagnosis (MCS vs. PVS); etiology of injury (TBI vs. non-TBI); time from injury to tDCS intervention (less than 3 months vs. more than 3 months); 1 session tDCS stimulation protocol vs. multi-sessions tDCS stimulation protocol; tDCS stimulation site (DLPFC vs. non-DLPFC). For all subgroup analyses, the same one-stage method was used for the analysis of the outcome. Additionally, treatment-by-subgroup interactions were tested by including multiplicative interaction terms in respective regression models. Subgroup analyses were again adjusted by prespecified covariates.

Moreover, we planned sensitivity analyses for trials with crossover randomized clinical trial study design, by excluding 2 trials with parallel randomized clinical trial study design. In another sensitivity analysis, we excluded one study which used 4mA current intensity protocol. All analyses were done with STATA, version 16.0. Forest plots were created by R, version 4.1.3 “forestplot” package.

## Result

### Study and Patient Characteristics

Our literature research identified 145 papers, 58 duplicated studies were removed, and 87 studies were screened by abstract and title. After full-text reviews and sought for IPD, 6 studies that met the eligibility criteria were excluded because no CRS-R score IPD were reported, and 8 trials [5-10, 18, 19] were eligible for inclusion in the meta-analysis: four done in Belgium, three in China and one in Italy. All 8 trials provided 181 individual patient data, among which 1 incomplete individual data was excluded from the analysis. We finally comprised 180 patients in the analyses. (**Figure 1**)

**Figure 1.**
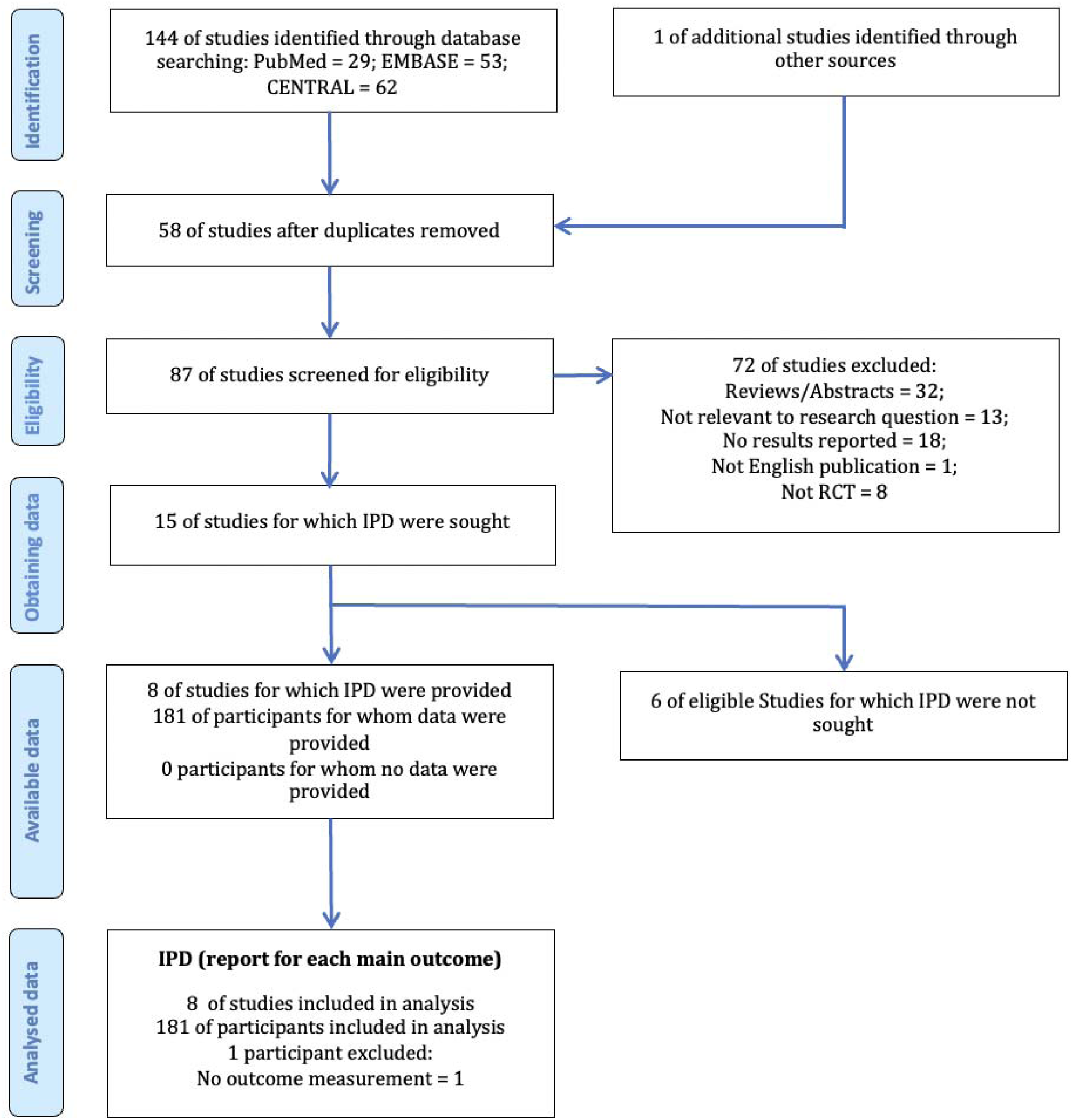
The PRISMA IPD flow diagram. *Abbreviations: IPD, Individual Patient Data; RCT, Randomized Clinical Trial; PRISMA, Preferred Reporting Items for Systematic Reviews and Meta-Analyses*.

Among the included trials, 41 participants of 2 trials were in parallel-RCT design, in which 18 participants were allocated to the sham-tDCS group, and 23 participants were allocated to the active-tDCS group; 139 participants of 6 trials were in crossover-RCT design, in which all 139 participants were allocated to both active-tDCS group and sham-tDCS group with a washout time interval between two groups. Because all crossover-RCT designed studies had reasonable washout periods and each crossover trial had pre and post-tDCS CRS-R scores in both active and sham groups, we comprised the data of all allocation for analyses. A sensitivity analysis was planned to account for this. In sum, we have 157 participants who received sham-tDCS and 162 who received active-tDCS.

Included studies are overviewed in **Table 1**. In the pooled study population, the mean (SD) age was 50 (17) years old, 68 (37.8%) patients were female, and 111 (61.7%) were male; the mean (SD) time from injury to tDCS intervention was 29 (56) months, 79 (43.9%) patients were caused by Traumatic Brain Injury (TBI), 101 (56.1%) were not; 127 (70.6%) patients were diagnosed as MCS at baseline, 52 (28.9%) were diagnosed as PVS. For stimulation protocol, 3 studies conducted the single session protocol, 2 with 5 sessions, 2 with 10 sessions, and 1with 20 sessions; 5 studies centered DLPFC as the stimulation site, 3 were not; the duration of the stimulation was 20 minus in each trial; all studies use a 2mA current intensity for stimulation except one (Martens used 4mA in stimulation), this study was accounted for a sensitivity analysis. Only one trial reported adverse effects, but the patient was excluded in that study, so all participants in our meta-analysis were well tolerant to the tDCS. Baseline characteristics and protocol characteristics of the 180 patients are shown in **Table 2**.

**Table 1.**
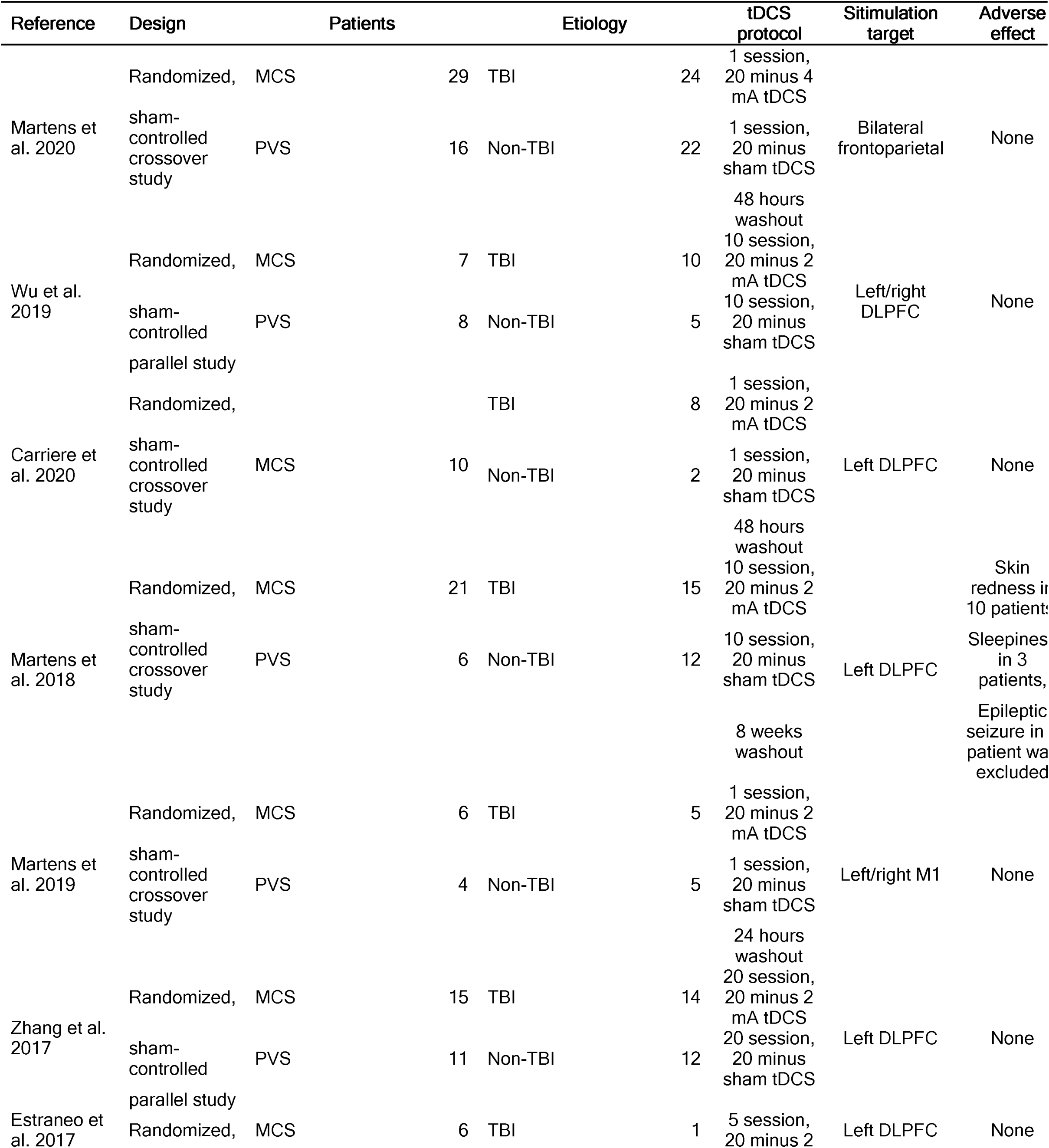
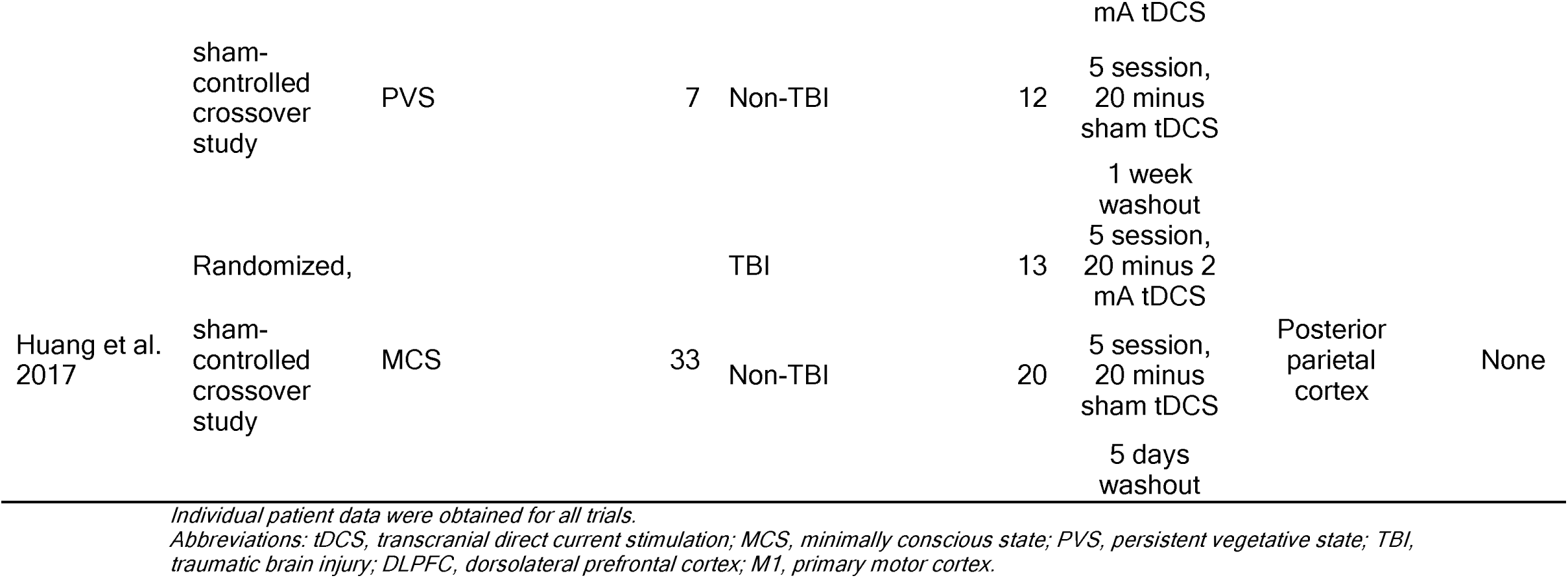
Overview of included studies.

**Table 2.** Overview of patient and protocol characteristics.

| Characteristics | value |
| --- | --- |
| Age, yr | 50±17 |
| Gender, n(%) |  |
| Male | 111 (61.7) |
| Female | 68 (37.8) |
| Etiology, n(%) |  |
| TBI | 79 (43.9) |
| Non-TBI | 101 (56.1) |
| Diagnosis, n(%) |  |
| MCS | 127 (70.6) |
| PVS | 52 (28.9) |
| Time from injury to tDCS, n(%) |  |
| 3 and less than 3 months | 48 (26.7) |
| More than 3 months | 132 (73.3) |
| Numbers of sessions, n(%) |  |
| 1 session | 66 (36.7) |
| more than 1 sessions | 114 (63.3) |
| Stimulation site, n(%) |  |
| DLPFC | 91 (50.6) |
| Non-DLPFC | 89 (49.4) |

*Abbreviations: TBI, traumatic brain injury; MCS, minimally conscious state; PVS, persistent vegetative state; DLPFC, dorsolateral prefrontal cortex; tDCS, transcranial direct current stimulation;*

Risk of bias were assessed for all trials. Results shows it has low to moderate risk of bias, which can be seen in **Appendix 2** in Supplementary data.

### Outcome

The main analysis of the outcome showed a significant result that the treatment effect (i.e., mean difference in the absolute CRS-R score change between pre and post-tDCS for treatment group versus control group, after adjusting for baseline characteristics) of 0.89 (95% CI, 0.17-1.61, *p* =0.015), with low heterogeneity across studies (I^2^ = 10.94%, τ^2^ = 0.53). (**Figure 2**) It indicates that compared with the control group, tDCS treatment could improve patients’ behavior performance measured by the CRS-R score, with a pooled mean difference of 0.89. Similar results could be obtained from the two-stage method, with a mean difference of 0.60 (95% CI, 0.23-0.96, *p* = 0.001). If not stated otherwise, all results are adjusted results.

**Figure 2.**
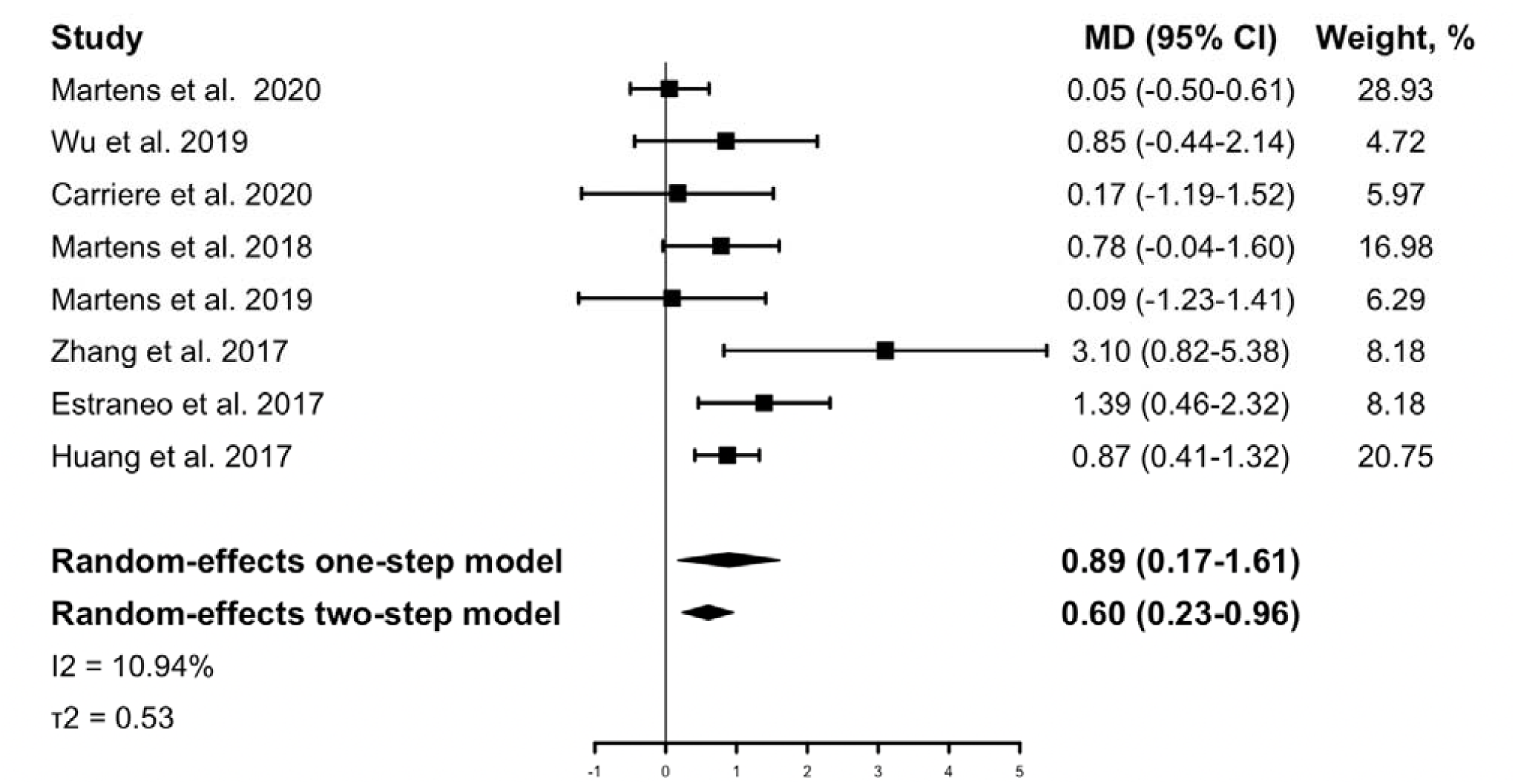
The IPD meta-analysis for all included studies: mean CRS-R score change between pre- and post-tDCS for the treatment group versus the control group. The Forest plot shows the positive MD both in the one-stage and two-stage meta-analysis method, meaning that active tDCS can significantly improve patients’ CRS-R scores than sham tDCS. The mean differences were adjusted for the following variables: age, sex, baseline CRS-R score, etiology of injury, time from injury to tDCS intervention. *Abbreviation: MD, mean difference; CRS-R, Coma Recovery Scale-Revised*.

### Subgroup analysis

In the subgroup analysis for the outcome (absolute CRS-R score change after active or sham tDCS), no evidence of heterogeneity of treatment effect was found across any of the following variables: etiology of injury (TBI vs. non-TBI), time from injury to tDCS intervention (less than 3 months vs. more than 3 months) or stimulation site (DLPFC vs. non-DLPFC). Among the patient who received only 1 session, no significant differences in treatment effect were observed between the active and sham group (−0.24, 95% CI [-1.97-1.47], *p* = 0.786); while in patients who received more than 1 sessions, significant improvements in CRS-R score could be observed (1.63, 95% CI [0.31-2.95], *p* = 0.016); but no statistically significant interaction was identified between session group and treatment effect (*P*_*interaction*_ = 0.091). Significant heterogeneity of treatment effect was observed for gender that male patients had a mean difference of 1.49 (95% CI, 0.18-2.80) in treatment effect, while female patients had a mean difference of 0.00 (95% CI, -1.40-1.39) in treatment effect.(*P*_*interaction*_ = 0.002). Treatment effect in patients’ baseline diagnosis also had a significant heterogeneity, in which patients were diagnosed as MCS had a mean difference of 1.37 (95% CI, 0.04 -2.70) in treatment effect while patients were diagnosed as PVS had a mean difference of -0.14 (95% CI, -1.63-1.36) in treatment effect. (*P*_*interaction*_ = 0.005). (**Figure 3**)

**Figure 3.**
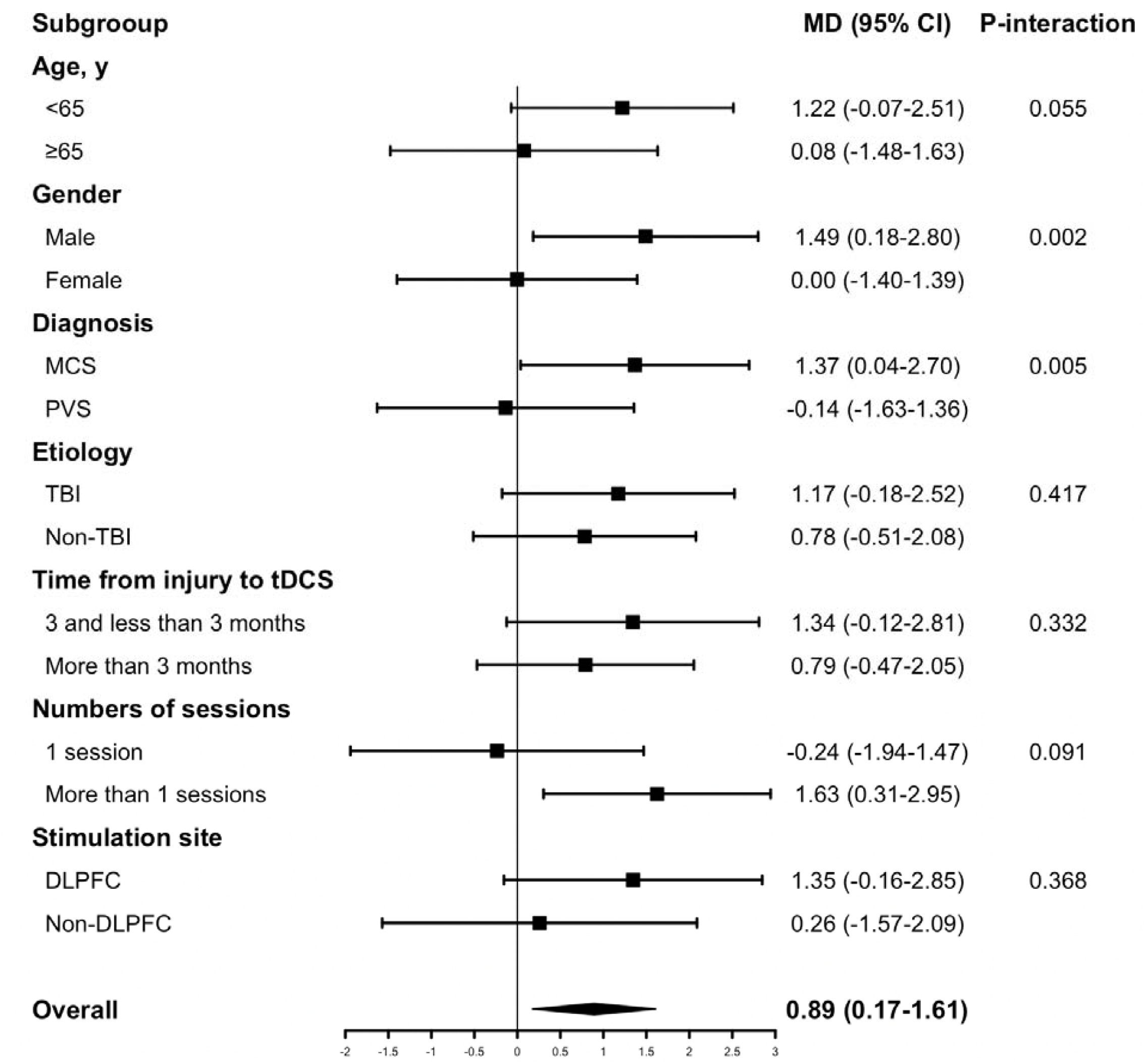
The subgroup analyses based on baseline characteristics and tDCS protocols. Subgroup analyses were adjusted by patients’ baseline covariates. *Abbreviations: MCS, minimally conscious state; PVS, persistent vegetative state; TBI, traumatic brain injury; DLPFC, dorsolateral prefrontal cortex*.

A sensitivity analysis excluding two trials that used parallel-RCT design did not show a substantial change in findings that tDCS significantly improved patients’ behavioral performance. (**Appendix 3**) Similar results could also be found in another sensitivity analysis in which the only trial that used a 4mA current intensity protocol was excluded. (**Appendix 4**)

## Discussion

The result of our meta-analysis of individual patient data from 8 randomized controlled trials showed that the tDCS treatment effect in patients with DoC is 0.89 (95% CI, 0.17-1.61), providing strong evidence that tDCS treatment is more effective at improving behavioral performance than control. This result was robust in all sensitivity analyses.

In previous knowledge, the behavioral treatment effects in DoC patients following tDCS were controversial. Three observational studies[20-22] and six clinical trials [10, 18, 19, 22-24]were consistent with our findings, whereas seven randomized clinical trials were confronted with ours that reported no significant treatment effect on behavior were observed. However, the sample size of these previous studies was too small, and in most of the studies, the patients’ baseline characteristics were unadjusted. It caused a limitation for these studies to properly assess the behavioral treatment effects of tDCS in DoC patients. To our knowledge, the present study is the first meta-analysis as well as of individual patient data about tDCS in the DoC. In our meta-analysis, individual data of 180 patients from a total of 8 studies[5-10, 18, 19] across different countries were used. After adjusting patients’ baseline characteristics, our results with a large sample size showed a more credible result than previous studies. Moreover, our meta-analysis of IPD allowed exploration of heterogeneity in treatment effect with subgroup analyses. In our subgroup analysis based on patient characteristics, geriatric patients (age ≥ 65 years) and patients under 65 had no heterogeneity of treatment effect (*P*_*interaction*_= 0.055). This finding should be interpreted with caution because the number of the older patients in our study was 36 (20%), so the estimate of treatment effect might be imprecise. Although no significant interaction was detected, there is a trend that patients under 65 have a more obvious treatment effect than patients over 65 (1.22 vs. 0.08). A larger sample size of older patients in future studies could generate more reliable results. Time from injury to tDCS intervention in patients less than 3 months and more than 3 months ruled out a possible heterogeneity of the treatment effect (*P*_*interaction*_ = 0.332), which indicates that DoC patients in both acute-subacute stage and chronic stage would benefit from tDCS treatment. No significant interaction between the etiology and treatment effect was evident. In the gender subgroup, there is a significant interaction between treatment effect and gender (*P*_*interaction*_= 0.002). However, no previous studies had reported such a difference before. One reason for this difference might be attributed to no such analysis in previous trials because of the relatively small sample size. A significant interaction between the diagnosis subgroup and treatment effect was also observed: the comparison between patients diagnosed with MCS and PVS yielded a *P*_*interaction*_ = 0.005. This result, consistent with two RCTs[11, 12], reported that the treatment effect was significant in MCS patients but not in PVS patients. In advance, in the current study, the MCS subgroup has an estimated absolute change in CRS-R score of 1.37, higher than the PVS subgroup. This might be helpful in guiding the clinical utilization of tDCS treatment in DoC patients. Admittedly, there might be some tDCS protocols that were effective in the PVS patients that need to be researched in the future.

For tDCS protocol, although no statistically significant interaction was identified between session subgroup and treatment effect, the benefit of tDCS in behavioral improvement was significant in patients who underwent multiple sessions tDCS protocol (1.63, 95% CI [0.31-2.95], *p* = 0.016), but did not reach statistically significant in patients who underwent one session tDCS stimulation protocol (−0.24, 95% CI [-1.97-1.47], *p* = 0.786). No significant difference was found within stimulation site subgroups.

In summary, our subgroup analysis identifies patients who are male or with an MCS diagnosis are most likely to respond to tDCS treatment. In addition, we found patients under the age of 65 or with multiple sessions of tDCS protocol might benefit from tDCS treatment. It is important to interpret these results cautiously.

### Limitations and Future Directions

There are some limitations to this study. In most individual studies, they not only assessed the behavioral effect of the tDCS measured via CRS-R score but also assessed the neurophysiological effect such as electroencephalogram (EEG) power spectra, EEG complexity, EEG connectivity analyses, and event-related potential (ERP) analyses. Some of them reported that the neurophysiological effect of tDCS could be found even though no behavioral changes were observed.[5, 6, 8-11, 25-27] Cavaliere et al. [20] reported that increased connectivity of the extrinsic control network was measured by functional Magnetic Resonance Imaging (fMRI) after tDCS. Thibaut et al.[21] found a transient increase of signs of consciousness after tDCS through fluorodeoxyglucose positron emission tomography (FDG-PET). However, these advanced measurements varied from study to study, making them difficult to be analyzed in the meta-analysis. Hence our study only analyzed the behavioral effect of the tDCS. In future studies, raw data of these neurophysiological assessments might be collected and processed with uniform methods.

Our meta-analysis identified patient and tDCS protocol characteristics associated with the improvement in CRS-R score. Due to the heterogeneity of included IPD, we have to transform these to create a uniform database that makes analysis possible. Thus, some information was lost by the transformation. For example, MCS was sub-stratified into MCS without language (MCS–) and MCS with language (MCS+)[28], a prognostic value might exist in the classification.[29] But in our analysis, patients were just divided into MSC and PVS, so the treatment effect between MCS+ and MCS-can not be analyzed. The same situation could also be seen in the etiology of the injury and stimulation site of tDCS. Future studies might focus on more detailed characteristics and their association with treatment effects. Finally, the interaction analyses in subgroup analysis may be limited to detecting heterogeneity in treatment outcomes, and the results should be interpreted cautiously. Despite the limitations, our study still provides strong evidence for utilizing tDCS in DoC.

## Conclusion

In this meta-analysis of patients with DoC, tDCS intervention could significantly improve patients’ clinical behavioral performance measured by the CRS-R score. We found that patients with certain baseline characteristics have a better improvement, while those with some characteristics do not. Future studies should focus on a more specific characteristic of patients who will respond to tDCS most and the tDCS protocol that would benefit patients most based on the mechanism of tDCS and neurophysiology.

## Supporting information

Spplemental Appendix 1

Spplemental Appendix 2, 3, and 4

## Data Availability

All data produced in the present study are available upon reasonable request to the authors.

## Author Contribution

**Zeyu Xu:** study conception, design, data acquisition, formal analysis, visualization and writing — original draft. **Tiantong Xia:** investigation, formal analysis. **Ruizhe Zheng:** data acquisition and writing — review and editing. **Xuehai Wu:** supervision, writing — review and editing.

**Zengxin Qi, Di Zang**, and **Zhe Wang:** data interpretation, revising the manuscript. All authors contributed to the article and approved the submitted version.

## Acknowledgement and Funding

We would like to acknowledge all the researchers who were willing to share their data that made this study possible, namely Géraldine Martens, Anna Estraneo, Min Wu, Wangshan Huang, Ye Zhang, and Manon Carrière. We also wish to thank the Shanghai Science and Technology Commission, **Xuehai Wu** is supported by the Shanghai Municipal Science and Technology Major Project (No.2018SHZDZX01).

