## Supplementary material for "Behavioral Effects in Disorders of Consciousness Following Transcranial Direct Current Stimulation: A Systematic Review and Individual Patient Data Meta-analysis of Randomized Clinical Trials": Spplemental Appendix 1

**Search Strategy**

Embase

Searched on April 7, 2022

No language, publication date or article type restrictions

1. 'disorder* of consciousness'

2. 'persistent vegetative state'/exp OR 'unresponsive wakefulness syndrome'/exp OR 'minimally conscious state'/exp OR 'states, transient vegetative' OR 'post-comatose unawareness states' OR 'state, post-comatose unawareness' OR 'persistent vegetative states' OR 'post-traumatic vegetative states' OR 'unawareness state, posttraumatic' OR 'minimally conscious state' OR 'state, post-traumatic unawareness' OR 'states, post-comatose unawareness' OR 'state, posttraumatic unawareness' OR 'states, posttraumatic unawareness' OR 'state, minimally conscious' OR 'postcomatose unawareness state' OR 'state, vegetative' OR 'permanent vegetative state' OR 'vegetative states, persistent' OR 'states, permanent vegetative' OR 'state, persistent unawareness' OR 'prolonged post-traumatic unawareness' OR 'minimally conscious states' OR 'transient vegetative states' OR 'state, permanent vegetative' OR 'states, vegetative' OR 'post comatose unawareness state' OR 'pvs' OR 'post-traumatic vegetative state' OR 'prolonged post-traumatic unawarenesses' OR 'vegetative state, transient' OR 'vegetative states, transient' OR 'states, post-traumatic vegetative' OR 'posttraumatic unawareness state' OR 'permanent vegetative states' OR 'unawareness state, postcomatose' OR 'vegetative states, permanent' OR 'states, persistent unawareness' OR 'vegetative state, permanent' OR 'vegetative state, post-traumatic' OR 'states, post-traumatic unawareness' OR 'unawareness, prolonged post-traumatic' OR 'states, minimally conscious' OR 'post traumatic vegetative state' OR 'unawareness states, post-comatose' OR 'post-comatose unawareness state' OR 'vegetative state' OR 'post-traumatic unawareness, prolonged' OR 'unawareness states, postcomatose' OR 'posttraumatic unawareness states' OR 'prolonged post traumatic unawareness' OR 'state, postcomatose unawareness' OR 'unawareness states, posttraumatic' OR 'unawareness state, post-comatose' OR 'post-traumatic unawareness states' OR 'persistent unawareness state' OR 'unawareness state, post-traumatic' OR 'state, post-traumatic vegetative' OR 'transient vegetative state' OR 'vegetative states, post-traumatic' OR 'persistent unawareness states' OR 'unawareness states, persistent' OR 'unawareness state, persistent' OR 'unawarenesses, prolonged post-traumatic' OR 'unawareness states, post-traumatic' OR 'post traumatic unawareness state' OR 'post-traumatic unawareness state' OR 'state, transient vegetative' OR 'states, postcomatose unawareness' OR 'state, persistent vegetative' OR 'states, persistent vegetative' OR 'post-traumatic unawarenesses, prolonged' OR 'postcomatose unawareness states' OR 'vegetative state, persistent' OR 'vegetative states' OR 'pvss'

3. 1 or 2

4. 'transcranial direct current stimulation'/exp

5. 'tdcs':ti,ab,kw OR 'cathodal stimulation transcranial direct current stimulation':ti,ab,kw OR 'cathodal stimulation tdcs':ti,ab,kw OR 'cathodal stimulation tdcss':ti,ab,kw OR 'stimulation tdcs, cathodal':ti,ab,kw OR 'stimulation tdcss, cathodal':ti,ab,kw OR 'tdcs, cathodal stimulation':ti,ab,kw OR 'tdcss, cathodal stimulation':ti,ab,kw OR 'transcranial random noise stimulation':ti,ab,kw OR 'transcranial alternating current stimulation':ti,ab,kw OR 'transcranial electrical stimulation':ti,ab,kw OR 'electrical stimulation, transcranial':ti,ab,kw OR 'electrical stimulations, transcranial':ti,ab,kw OR 'stimulation, transcranial electrical':ti,ab,kw OR 'stimulations, transcranial electrical':ti,ab,kw OR 'transcranial electrical stimulations':ti,ab,kw OR 'anodal stimulation transcranial direct current stimulation':ti,ab,kw OR 'anodal stimulation tdcs':ti,ab,kw OR 'anodal stimulation tdcss':ti,ab,kw OR 'stimulation tdcs, anodal':ti,ab,kw OR 'stimulation tdcss, anodal':ti,ab,kw OR 'tdcs, anodal stimulation':ti,ab,kw OR 'tdcss, anodal stimulation':ti,ab,kw OR 'repetitive transcranial electrical stimulation'

6. 4 or 5

7. 'clinical':ti,ab AND 'trial':ti,ab OR 'clinical trial'/exp OR random* OR 'drug therapy':lnk

8. 3 and 6 and 7	53

Pubmed

Searched on April 7, 2022

No language, publication date or article type restrictions

((((("Persistent Vegetative State"[Mesh]) OR (PVS (Persistent Vegetative State) OR PVSs (Persistent Vegetative State) OR Vegetative State, Persistent OR Persistent Vegetative States OR State, Persistent Vegetative OR States, Persistent Vegetative OR Vegetative States, Persistent OR Persistent Unawareness State OR Persistent Unawareness States OR State, Persistent Unawareness OR States, Persistent Unawareness OR Unawareness State, Persistent OR Unawareness States, Persistent OR Permanent Vegetative State OR Permanent Vegetative States OR State, Permanent Vegetative OR States, Permanent Vegetative OR Vegetative State, Permanent OR Vegetative States, Permanent OR Vegetative State OR State, Vegetative OR States, Vegetative OR Vegetative States OR Post-Traumatic Vegetative State OR Post Traumatic Vegetative State OR Post-Traumatic Vegetative States OR State, Post-Traumatic Vegetative OR States, Post-Traumatic Vegetative OR Vegetative State, Post-Traumatic OR Vegetative States, Post-Traumatic OR Posttraumatic Unawareness State OR Posttraumatic Unawareness States OR State, Posttraumatic Unawareness OR States, Posttraumatic Unawareness OR Unawareness State, Posttraumatic OR Unawareness States, Posttraumatic OR Post-Traumatic Unawareness State OR Post Traumatic Unawareness State OR Post-Traumatic Unawareness States OR State, Post-Traumatic Unawareness OR States, Post-Traumatic Unawareness OR Unawareness State, Post-Traumatic OR Unawareness States, Post-Traumatic OR Prolonged Post-Traumatic Unawareness OR Post-Traumatic Unawareness, Prolonged OR Post-Traumatic Unawarenesses, Prolonged OR Prolonged Post Traumatic Unawareness OR Prolonged Post-Traumatic Unawarenesses OR Unawareness, Prolonged Post-Traumatic OR Unawarenesses, Prolonged Post-Traumatic OR Transient Vegetative State OR State, Transient Vegetative OR States, Transient Vegetative OR Transient Vegetative States OR Vegetative State, Transient OR Vegetative States, Transient OR Minimally Conscious State OR Minimally Conscious States OR State, Minimally Conscious OR States, Minimally Conscious OR Post-Comatose Unawareness State OR Post Comatose Unawareness State OR Post-Comatose Unawareness States OR State, Post-Comatose Unawareness OR States, Post-Comatose Unawareness OR Unawareness State, Post-Comatose OR Unawareness States, Post-Comatose OR Postcomatose Unawareness State OR Postcomatose Unawareness States OR State, Postcomatose Unawareness OR States, Postcomatose Unawareness OR Unawareness State, Postcomatose OR Unawareness States, Postcomatose)) OR (((unresponsive wakefulness syndrome) OR (disorder of consciousness[Title/Abstract])) OR (disorders of consciousness[Title/Abstract])))) AND (("Transcranial Direct Current Stimulation"[Mesh]) OR ((tDCS[Title/Abstract]) OR (Cathodal Stimulation Transcranial Direct Current Stimulation[Title/Abstract]) OR (Cathodal Stimulation tDCS[Title/Abstract]) OR (Cathodal Stimulation tDCSs[Title/Abstract]) OR (Stimulation tDCS, Cathodal[Title/Abstract]) OR (Stimulation tDCSs, Cathodal[Title/Abstract]) OR (tDCS, Cathodal Stimulation[Title/Abstract]) OR (tDCSs, Cathodal Stimulation[Title/Abstract]) OR (Transcranial Random Noise Stimulation[Title/Abstract]) OR (Transcranial Alternating Current Stimulation[Title/Abstract]) OR (Transcranial Electrical Stimulation[Title/Abstract]) OR (Electrical Stimulation, Transcranial[Title/Abstract]) OR (Electrical Stimulations, Transcranial[Title/Abstract]) OR (Stimulation, Transcranial Electrical[Title/Abstract]) OR (Stimulations, Transcranial Electrical[Title/Abstract]) OR (Transcranial Electrical Stimulations[Title/Abstract]) OR (Anodal Stimulation Transcranial Direct Current Stimulation[Title/Abstract]) OR (Anodal Stimulation tDCS[Title/Abstract]) OR (Anodal Stimulation tDCSs[Title/Abstract]) OR (Stimulation tDCS, Anodal[Title/Abstract]) OR (Stimulation tDCSs, Anodal[Title/Abstract]) OR (tDCS, Anodal Stimulation[Title/Abstract]) OR (tDCSs, Anodal Stimulation[Title/Abstract]) OR (Repetitive Transcranial Electrical Stimulation[Title/Abstract])))) AND ((clinical[tiab] AND trial[tiab]) OR "clinical trials as topic"[mesh] OR "clinical trial"[pt] OR random*[tiab] OR "random allocation"[mesh] OR "therapeutic use"[sh])	29

Cochrane CENTRAL

Searched on April 7, 2022

No language, publication date or article type restrictions

1. (States, Transient Vegetative) OR (Post-Comatose Unawareness States) OR (State, Post-Comatose Unawareness) OR (Persistent Vegetative States) OR (Post-Traumatic Vegetative States) OR (Unawareness State, Posttraumatic) OR (Minimally Conscious State) OR (State, Post-Traumatic Unawareness) OR (States, Post-Comatose Unawareness) OR (State, Posttraumatic Unawareness) OR (States, Posttraumatic Unawareness) OR (State, Minimally Conscious) OR (Postcomatose Unawareness State) OR (State, Vegetative) OR (Permanent Vegetative State) OR (Vegetative States, Persistent) OR (States, Permanent Vegetative) OR (State, Persistent Unawareness) OR (Prolonged Post-Traumatic Unawareness) OR (Minimally Conscious States) OR (Transient Vegetative States) OR (State, Permanent Vegetative) OR (States, Vegetative) OR (Post Comatose Unawareness State) OR (Post-Traumatic Vegetative State) OR (Prolonged Post-Traumatic Unawarenesses) OR (Vegetative State, Transient) OR (Vegetative States, Transient) OR (States, Post-Traumatic Vegetative) OR (Posttraumatic Unawareness State) OR (Permanent Vegetative States) OR (Unawareness State, Postcomatose) OR (Vegetative States, Permanent) OR (States, Persistent Unawareness) OR (Vegetative State, Permanent) OR (Vegetative State, Post-Traumatic) OR (States, Post-Traumatic Unawareness) OR (Unawareness, Prolonged Post-Traumatic) OR (States, Minimally Conscious) OR (Post Traumatic Vegetative State) OR (Unawareness States, Post-Comatose) OR (Post-Comatose Unawareness State) OR (Vegetative State) OR (Post-Traumatic Unawareness, Prolonged) OR (Unawareness States, Postcomatose) OR (Posttraumatic Unawareness States) OR (Prolonged Post Traumatic Unawareness) OR (State, Postcomatose Unawareness) OR (Unawareness States, Posttraumatic) OR (Unawareness State, Post-Comatose) OR (Post-Traumatic Unawareness States) OR (Persistent Unawareness State) OR (Unawareness State, Post-Traumatic) OR (State, Post-Traumatic Vegetative) OR (Transient Vegetative State) OR (Vegetative States, Post-Traumatic) OR (Persistent Unawareness States) OR (Unawareness States, Persistent) OR (Unawareness State, Persistent) OR (Unawarenesses, Prolonged Post-Traumatic) OR (Unawareness States, Post-Traumatic) OR (Post Traumatic Unawareness State) OR (Post-Traumatic Unawareness State) OR (State, Transient Vegetative) OR (States, Postcomatose Unawareness) OR (State, Persistent Vegetative) OR (States, Persistent Vegetative) OR (Post-Traumatic Unawarenesses, Prolonged) OR (Postcomatose Unawareness States) OR (Vegetative State, Persistent) OR (Vegetative States) OR (disorder* of consciousness) OR (unresponsive wakefulness syndrome)

2. MeSH descriptor: [Persistent Vegetative State] explode all trees

3. 1 OR 2

4. MeSH descriptor: [Transcranial Direct Current Stimulation] explode all trees

5. (tDCS):ti,ab,kw OR (Cathodal Stimulation Transcranial Direct Current Stimulation):ti,ab,kw OR (Cathodal Stimulation tDCS):ti,ab,kw OR (Cathodal Stimulation tDCSs):ti,ab,kw OR (Stimulation tDCS, Cathodal):ti,ab,kw OR (Stimulation tDCSs, Cathodal):ti,ab,kw OR (tDCS, Cathodal Stimulation):ti,ab,kw OR (tDCSs, Cathodal Stimulation):ti,ab,kw OR (Transcranial Random Noise Stimulation):ti,ab,kw OR (Transcranial Alternating Current Stimulation):ti,ab,kw OR (Transcranial Electrical Stimulation):ti,ab,kw OR (Electrical Stimulation, Transcranial):ti,ab,kw OR (Electrical Stimulations, Transcranial):ti,ab,kw OR (Stimulation, Transcranial Electrical):ti,ab,kw OR (Stimulations, Transcranial Electrical):ti,ab,kw OR (Transcranial Electrical Stimulations):ti,ab,kw OR (Anodal Stimulation Transcranial Direct Current Stimulation):ti,ab,kw OR (Anodal Stimulation tDCS):ti,ab,kw OR (Anodal Stimulation tDCSs):ti,ab,kw OR (Stimulation tDCS, Anodal):ti,ab,kw OR (Stimulation tDCSs, Anodal):ti,ab,kw OR (tDCS, Anodal Stimulation):ti,ab,kw OR (tDCSs, Anodal Stimulation):ti,ab,kw OR (Repetitive Transcranial Electrical Stimulation)

6. 4 or 5

7. 3 and 6	62 in CENTRAL
