## Supplementary material for "Behavioral Effects in Disorders of Consciousness Following Transcranial Direct Current Stimulation: A Systematic Review and Individual Patient Data Meta-analysis of Randomized Clinical Trials": Spplemental Appendix 2, 3, and 4

**Appendix 2.** Risk of bias of each included trial with the updated version of the Risk of Bias Tool developed by Cochrane

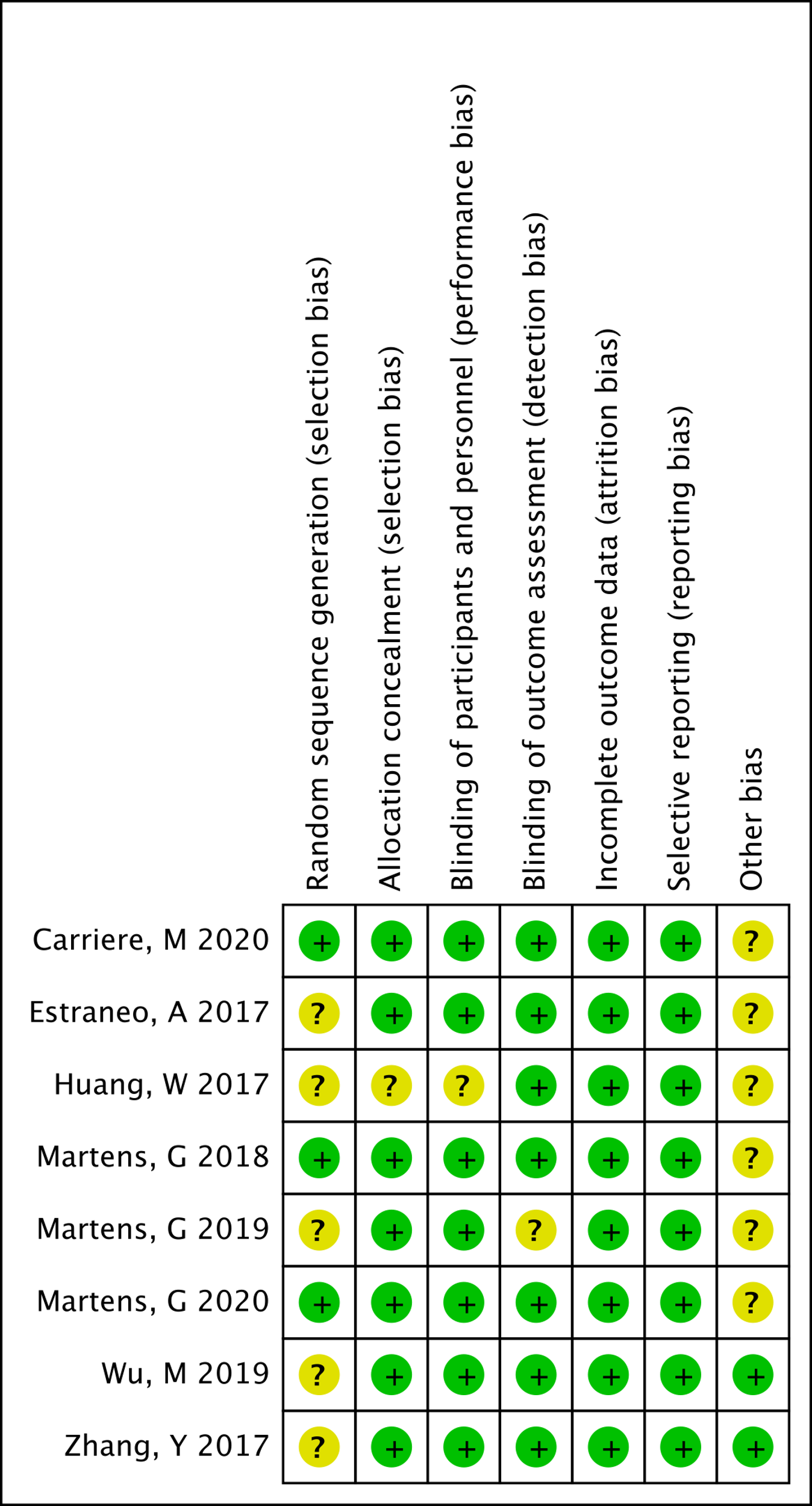

**Appendix 3.** Sensitivity analysis—crossover design randomized clinical trials only (Wu et al. 2019 and Zhang et al. 2017 were excluded)

|  | **Crude MD (95% CI)** | **Adjusted MD (95% CI)** | **P-value** |
| --- | --- | --- | --- |
| Treatment effect | 0.44 (0.02-0.88) | 0.44 (0.02-0.87) | 0.039 |

Adjusted the following variables: age, sex, baseline CRS-R score, etiology of injury, time from injury to tDCS intervention.

All analyses were performed with a one-stage model with random effects.

P-value was the significance of the adjusted one-stage IPD meta-analysis.

The treatment effect was defined as the mean CRS-R score change between pre- and post-tDCS for the treatment group versus the control group.

Abbreviation: MD, mean difference; CRS-R, Coma Recovery Scale-Revised; tDCS, transcranial direct current stimulation; IPD, Individual Patient Data.

**Appendix 4.** Sensitivity analysis—2 mA tDCS protocol only (Martens et al. 2020 was excluded)

|  | **Crude MD (95% CI)** | **Adjusted MD (95% CI)** | **P-value** |
| --- | --- | --- | --- |
| Treatment effect | 1.15 (0.45-1.85) | 1.10 (0.53-1.67) | <0.001 |

Adjusted the following variables: age, sex, baseline CRS-R score, etiology of injury, time from injury to tDCS intervention.

All analyses were performed with a one-stage model with random effects.

P-value was the significance of the adjusted one-stage IPD meta-analysis.

The treatment effect was defined as the mean CRS-R score change between pre- and post-tDCS for the treatment group versus the control group.

Abbreviation: MD, mean difference; CRS-R, Coma Recovery Scale-Revised; tDCS, transcranial direct current stimulation; IPD, Individual Patient Data.
